# The burden trend and risk factors of multiple myeloma from 1990 to 2021: a systematic analysis for the Global Burden of Disease Study 2021

**DOI:** 10.1101/2025.04.20.25326120

**Authors:** Chang Wang, Yan Guo, Ning Wang, Qinduo Ren, Yuan Xu, Songlin Lu, Yuping Zou, Ruilin Li, Jiacheng Wang, Linna Yuan, Wei She, Hongsheng Tian, Shuo Bi, Xuying Guo, Ye Ma, Hongmei Sun, Chen Zhang, Yu Dong, Zhenwei Shang, Hongchao Lv, Wenhua Lv, Mingming Zhang, Yongshuai Jiang

## Abstract

**IMPORTANCE:** MM was an incurable clonal plasma cell malignancy with substantial prevalence and incidence, which had resulted in a significant burden of disease. Our study aimed to evaluate the global, regional and national burden of MM by sex, age, and SDI from 1990 to 2021, along with associated risk factors and predicted temporal trends.

**OBJECTIVE:** To assess the global burden, epidemiological trends, and risk factors of MM and project its incidence rate by 2050.

**Design:** This is a systematic analysis of the global burden of MM from 1990 to 2021.

**Setting:** The study primarily analyzed the prevalence, incidence, mortality, and disability-adjusted life years (DALYs) of MM from Global Burden of Diseases, Injuries and Risk Factors Study (GBD) 2021, by region, age, sex, and SDI.

**Participants:** The study builds on findings from the GBD 2021.

**Exposure:** Multiple myeloma.

**Main Outcomes and Measures:** The Joinpoint regression was used to describe the trends. Two-sample Mendelian randomization (MR) analysis was conducted to identify the causal relationship between weight-related risk factors and MM. The Bayesian age-period-cohort (BAPC) model was used to predict the future trend.

**Results:** In 2021, the age-standardized prevalence rate (ASPR) of MM was 4.65 (95% UI: 4.21 to 5.01) per 100,000 people, and the age-standardized mortality rate (ASMR) was 1.40 (95% UI: 1.24 to 1.52) per 100,000 people globally. The burden of MM was the highest in the high SDI region, with Monaco and Bahamas reporting the highest age-standardized incidence rate (ASIR), age-standardized DALY rate (ASDR), and ASMR in 2021. Simultaneously, ASRs peaked in the elderly population and were higher in males than in females. GBD 2021 reported high body mass index (BMI) as a risk factor for MM which was further confirmed by two-sample MR analysis. Additionally, BMI, waist circumference, obesity class 1, obesity class 2, overweight and hip circumference were also risk factors for MM. By 2050, the global incidence rate was predicted to decrease slightly but increase within the age group of 20-69.

**Conclusions and Relevance:** This study found that MM remained a significant public health issue, particularly in high SDI regions and among the elderly. A causal relationship exists between MM and body weight, indicating the need for targeted prevention and intervention strategies in high-risk regions and populations.

## Introduction

Multiple myeloma (MM), an incurable cancer of the immunoglobulin-producing plasma cells, was the second most hematological disease, which mainly developed in elderly population.^1,2^ The common clinical manifestations of MM were bone pain, anemia, renal function damage, hypercalcemia and infection which may severely reduce the quality of life and the expectancy of patients.^3,4^ In 2018, the new cases of MM were almost 1.5 times higher than that of deaths.^5^ The incidence had been reported increased by 136% from 1990 to 2019.^6^ All of these indicated that the global disease burden of MM was serious. Therefore, a comprehensive and systematic investigation into the disease burden and epidemiological trends of MM was imperative.

Existing studies on the global disease burden of MM remained insufficient. Certain studies on the burden of MM were limited to particular countries or regions, such as Neves et al. assessed the burden and cost of MM using the data provided by national data sources in Portugal,^7^ Wang et al. analyzed the gender, age, and area differences in the prevalence and incidence rate of MM based on the national medical insurance database in mainland China.^8^ Meanwhile, the Global Burden of Diseases, Injuries and Risk Factors Study (GBD) also provided a comprehensive and systematic framework to quantify the burden of MM. Using GBD 2016, Cowan et al. identified that the incidence of MM increased significantly from 1990 to 2016, predominantly in middle socio-demographic index (SDI) countries and East Asia.^9^ Based on the quality of care index calculated using GBD 2019, Geng et al. also pointed out that countries and regions with higher levels of socioeconomic development tend to have better quality of care for MM, while women and elderly patients were disadvantaged in terms of care.^10^ However, the date they utilized were relatively old. It was essential to conduct a comprehensive and lasted research on the burden of MM.

Compared to previous GBD, GBD 2021 contained more recent data and broader coverage of disease and risk factors. Based on the prevalence rate, incidence rate, disability-adjusted life-years (DALYs) and mortality rate of MM from GBD 2021, we analyzed the global, regional and national burden of MM. In addition, we explored the differences in the burden of MM across age, sex, and SDI. Moreover, the potential causal relationships between risk factors and MM were analyzed in detail and the incidence rate were predicted from 2022 to 2050. This systematic study could contribute to the development of effective prevention and treatment strategies.

## Methods

### Data source

Data for the disease burden of MM were from GBD 2021 (https://ghdx.healthdata.org/gbd-2021), which captured 288 causes of death, 371 diseases and injuries, and 88 risk factors in 204 countries and territories, by age and sex, from 1990 to 2021.^11^ From GBD 2021, we obtained the estimated incidence rate, prevalence rate, DALY and mortality rate of MM with 95% uncertainly interval (UI). (Details are provided in the supplemental materials p1)

### Age-standardized rate

The age-standardized rate (ASR) was the weighted arithmetic mean of age-specific rates per 100,000 people, with weights corresponding to the ratio of people in the standard population’s respective age groups. The ASR could be used to assess the global burden of MM across different countries and regions in different years, in terms of age-standardized prevalence rate (ASPR), age-standardized incidence rate (ASIR), age-standardized DALY rate (ASDR) and age-standardized mortality rate (ASMR). (supplemental materials p1)

### Average annual percent change

Average annual percent change (AAPC) was a summary measure of the trend over a pre-specified fixed interval, calculated as a weighted average of the annual percent changes from the Joinpoint model, with the weights equal to the length of the annual percent change interval.^12^ In our study, the AAPCs of ASRs along with 95% confidence interval (CI) were computed by Joinpoint regression 5.0 software. Based on the AAPC, trends of ASRs were identified as increasing (AAPC > 0) and decreasing (AAPC < 0).

### Socio-demographic Index

Socio-demographic index (SDI) was a composite indicator of development status strongly correlated with healthy outcomes. It was the geometric mean total fertility rate of under the age of 25, mean education for those ages 15 and older, and lag distributed income per capita, ranging from 0 to 100.^13^ According to the SDI, the 204 countries and territories were categorized into high SDI, high-middle SDI, middle SDI, low-middle SDI, and low SDI regions.^14^ Pearson correlation analysis was conducted to examine the relationship between SDI and ASR, as well as AAPC, across different countries.

### Two-sample Mendelian randomization

Mendelian randomization (MR) was a statistical method to analyze the causal relationship between an exposure and an outcome.^15^ In this study, we employed two-sample MR method to assess potential causal relationships between weight-related risk factors and MM, with the inverse variance weighted method serving as the primary analytical criterion to estimate the causal effects. The genome-wide association study (GWAS) summary data of MM (finn-b-I9_VHD, 2021) and risk factors were obtained from the OpenGWAS project. (supplemental materials p1)

### Bayesian age-period-cohort analysis

The Bayesian age-period-cohort (BAPC) analysis model was conducted based on the assumption that the effects of age, period, and cohort adjacent in time are similar, which could be used to project posterior incidence rates ^16^. Using BAPC (Version 0.0.36) and INLA (Version 24.2.9) packages in the R program (Version 4.4.1) for analysis,^17,18^ we embarked on forecasting the MM burden for the period 2022 to 2050.

## Results

### Global Burden of MM from 1990 to 2021

In 2021, it was estimated that there were 394,481.73 (95% UI: 355,593.17 to 425,503.10) prevalent cases of MM globally, which had increased by 218.20% since 1990 (Table1). Also, the number of incident cases, deaths and DALYs increased by 167.02%, 144.61% and 131.23%, respectively. From 1990 to 2021, the ASRs of MM also experienced a steady rise each year. The ASPR increased from 3.13 (95% UI: 2.96 to 3.30) to 4.55 per 100,000 population with an AAPC of 1.21 (95% CI: 1.17 to 1.24). Compared to 1.47 in 1990, the ASIR reached 1.74 per 100,000 population in 2021, with an AAPC of 0.55. Moreover, the AAPCs of ASDR and ASMR were 0.18 and 0.20, respectively. Overall, the global burden of MM had become gradually severe.

**Table 1.**
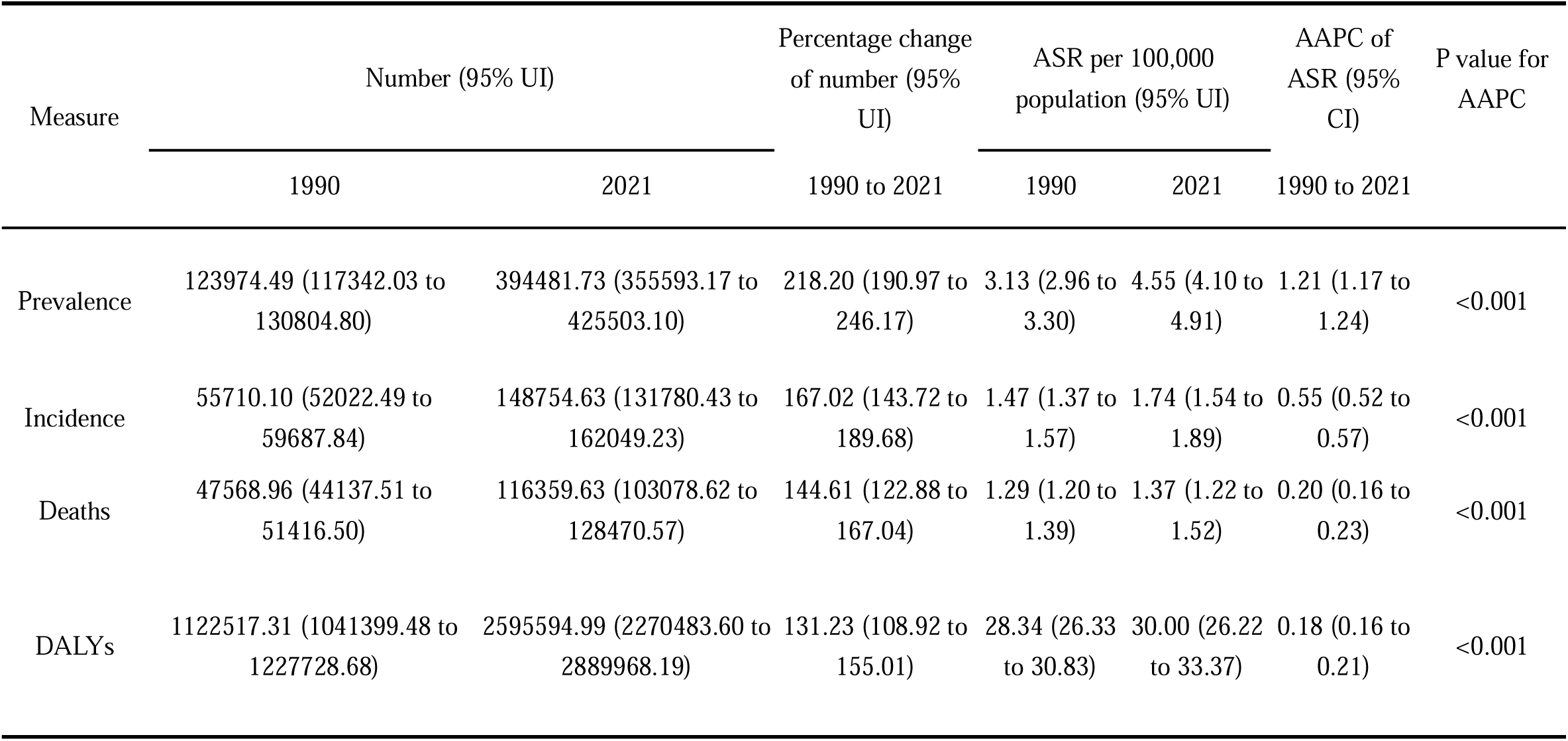
Global prevalent cases, incident cases, deaths, DALYs and ASRs with AAPC of MM in 1990 and 2021.

### Burden of MM in 21 regions from 1990 to 2021

In 2021, Australasia showed the heaviest disease burden with the highest ASPR (23.18 per 100,000 population), ASIR (5.48 per 100,000 population), ASMR (2.89 per 100,000 population), and ASDR (60.65 per 100,000 population) across all regions (Table S1). Meanwhile, East Asia presented the greatest upward trends in disease burden with the highest AAPCs of 6.03 for ASPR, 4.49 for ASIR, 3.74 for ASMR, and 3.71 for ASDR. Notably, although High-income North America had relatively high ASMR (2.81 per 100,000 population) and ASDR (57.28 per 100,000 population), it exhibited a decreasing trend with AAPCs of -0.49 for ASMR and -0.84 for ASDR. Among all of the regions, the lightest disease burden was observed in Oceania, with the lowest ASRs.

### Burden of MM in 204 countries and territories from 1990 to 2021

In 2021, the burden of MM varied across different countries and territories. The highest ASPR were observed in New Zealand (25.35 per 100,000 population), followed by Monaco (23.46 per 100,000 population) and Australia (22.75 per 100,000 population) (Figure 1, Table S2). Meanwhile, Monaco and Bahamas also showed relatively high ASIR, ASMR and ASDR, indicating a severe disease burden (Figure S1, Table S3-5). From 1990 to 2021, Mauritius exhibited the greatest increasing trends with the highest AAPCs of 7.15 for ASPR, 6.54 for ASIR, 6.28 for ASMR, and 6.33 for ASDR (Figure S2-5, Table S2-5). Additionally, China ranked second with an AAPC of 6.43 for ASPR and Georgia had the second highest AAPCs of 5.56 for ASIR, 5.50 for ASMR, and 6.33 for ASDR. Conversely, San Marino exhibited a relatively modest increase in the burden of MM with lower AAPCs for all ASRs.

**Figure 1.**
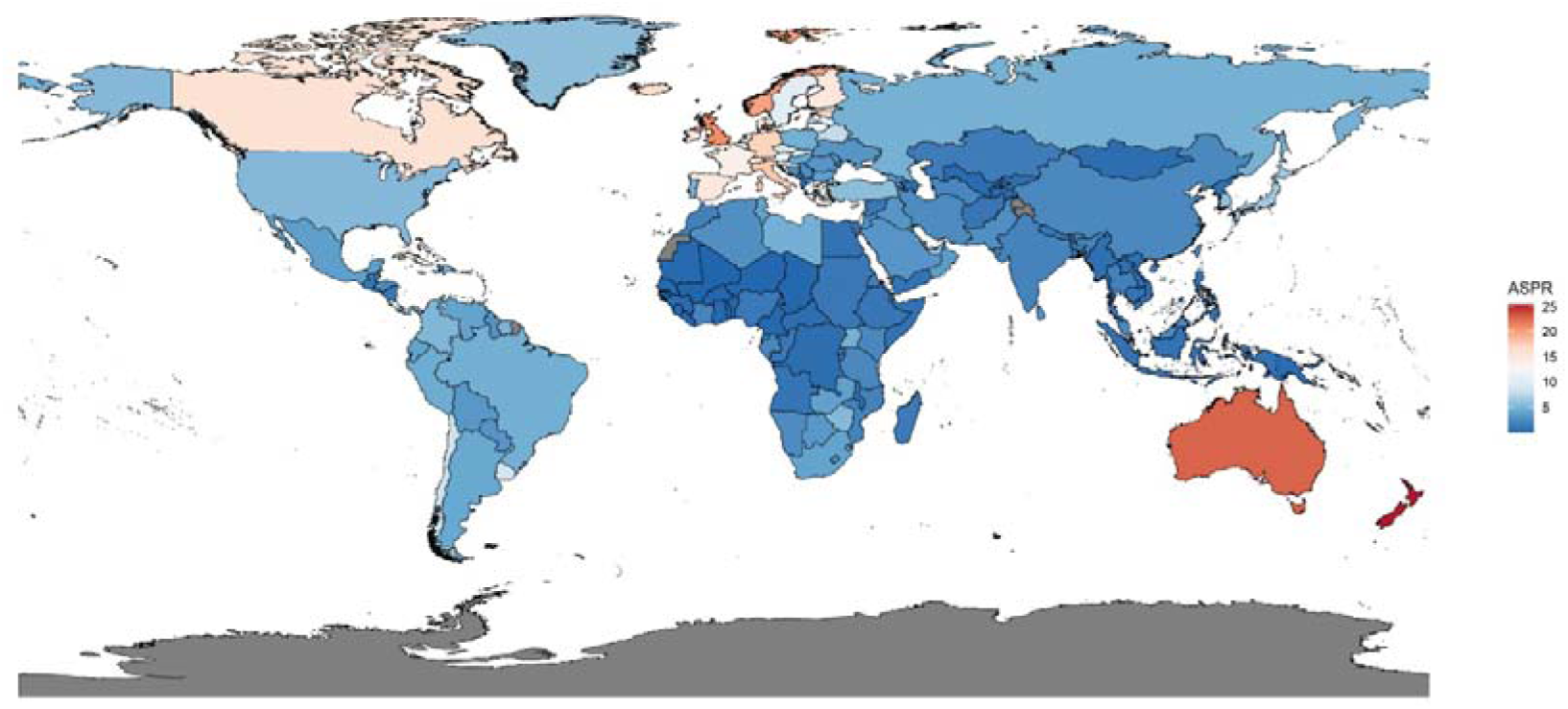
ASPR of MM in 204 countries and territories in 2021.

### The impact of age and sex pattern on global burden of MM

In 2021, the number of prevalent cases, incident cases, and deaths accounted for more than 70% of the total in the age group of 60-89 and peaked in the age group of 70-74, indicating that the global burden of MM was heaviest in the elderly population. Meanwhile, the global mortality rate increased with age, reaching 39.68 per 100,000 population for males and 24.81 per 100,000 population for females, both in the 95-years-and-over age group (Figure 2A). Similarly, the incidence and DALY rate peaked in the age group of 90-94, and prevalence rate peaked in the age group of 85-89 for both sexes (Figure 2B-D). Furthermore, males experienced a heavier MM burden. In the age group of 20-89, the number of incident cases in males was higher than in females with the largest gap of 3261.40 observed in the age group of 70-74. This phenomenon was similar in prevalent cases, deaths, and DALYs. Moreover, the t-tests on ASRs for sex differences revealed that the ASIR (*P* = 0.0039), ASMR (*P* = 0.0003), and ASDR (*P* = 0.0004) were all significantly higher in males compared to females (Figure S6).

**Figure 2.**
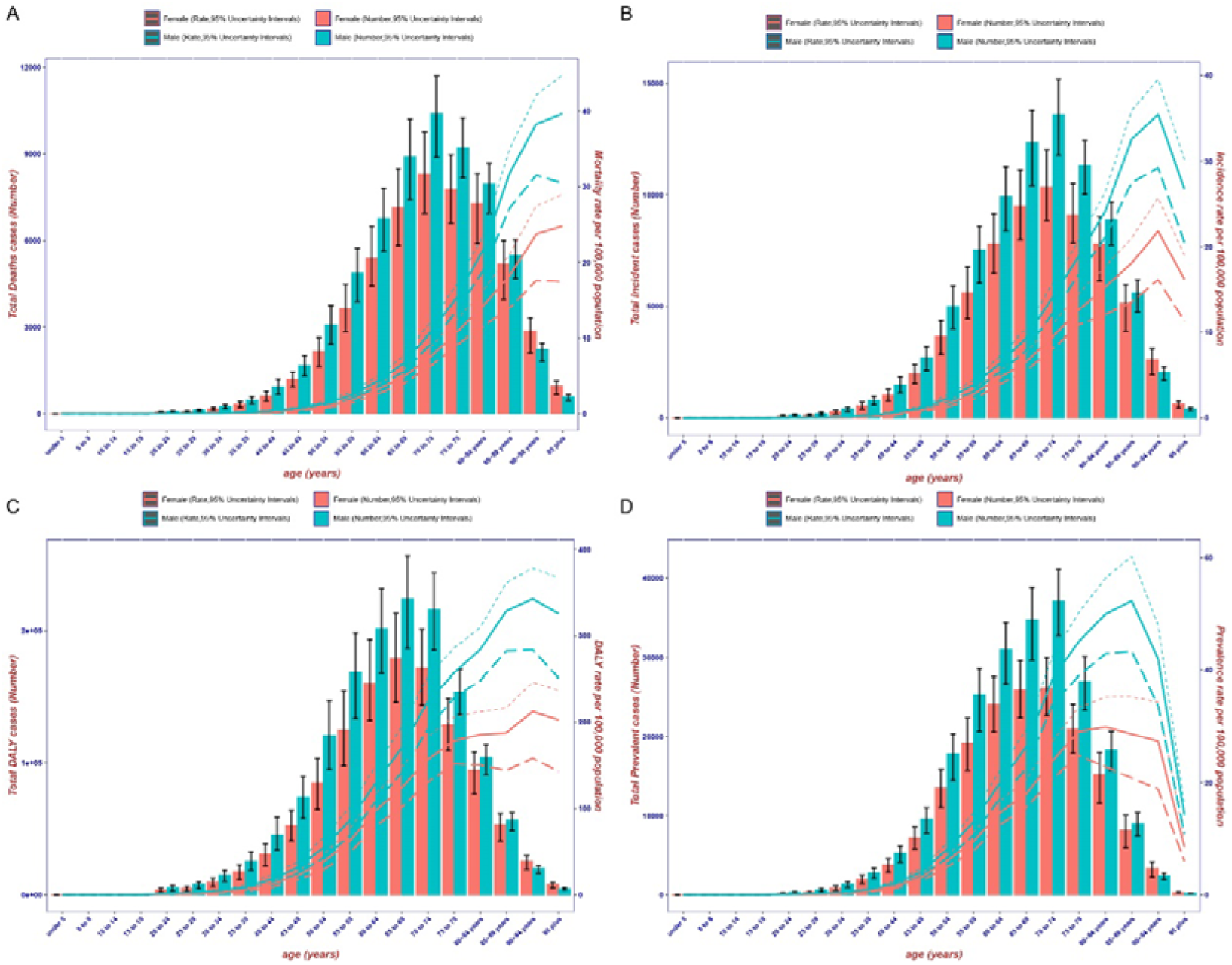
The global number of cases and ASRs of MM by age and sex in 2021. (A) Deaths; (B) Incidence; (C) DALYs; (D) Prevalence. The upper and lower dashed lines represent 95% of the upper and lower UI, respectively.

### The impact of SDI on burden of MM

From 1990 to 2021, high SDI regions experienced the most severe disease burden, exhibiting the highest ASRs, far exceeding the global level (Figure 3). Meanwhile, the ASPR and ASIR in high SDI regions increased with AAPCs of 1.03 and 0.16, respectively, while both the ASMR and ASDR decreased. Although ASRs in middle SDI, low-middle, and low SDI regions were below the global level, they all showed an upward trend. Among these five different SDI regions, the middle SDI regions had the largest AAPCs for ASRs (ASPR: 3.25; ASIR:2.35; ASMR: 1.99; ASDR: 1.91). In addition, the burden of MM was heavier in males than in females, with males exhibited higher ASRs than females within each SDI region. Furthermore, Pearson correlation analysis revealed a significant positive correlation between ASRs and SDI, while the relationship between AAPCs for all ASRs and SDI followed an inverted U-shaped curve (Figure S7).

**Figure 3.**
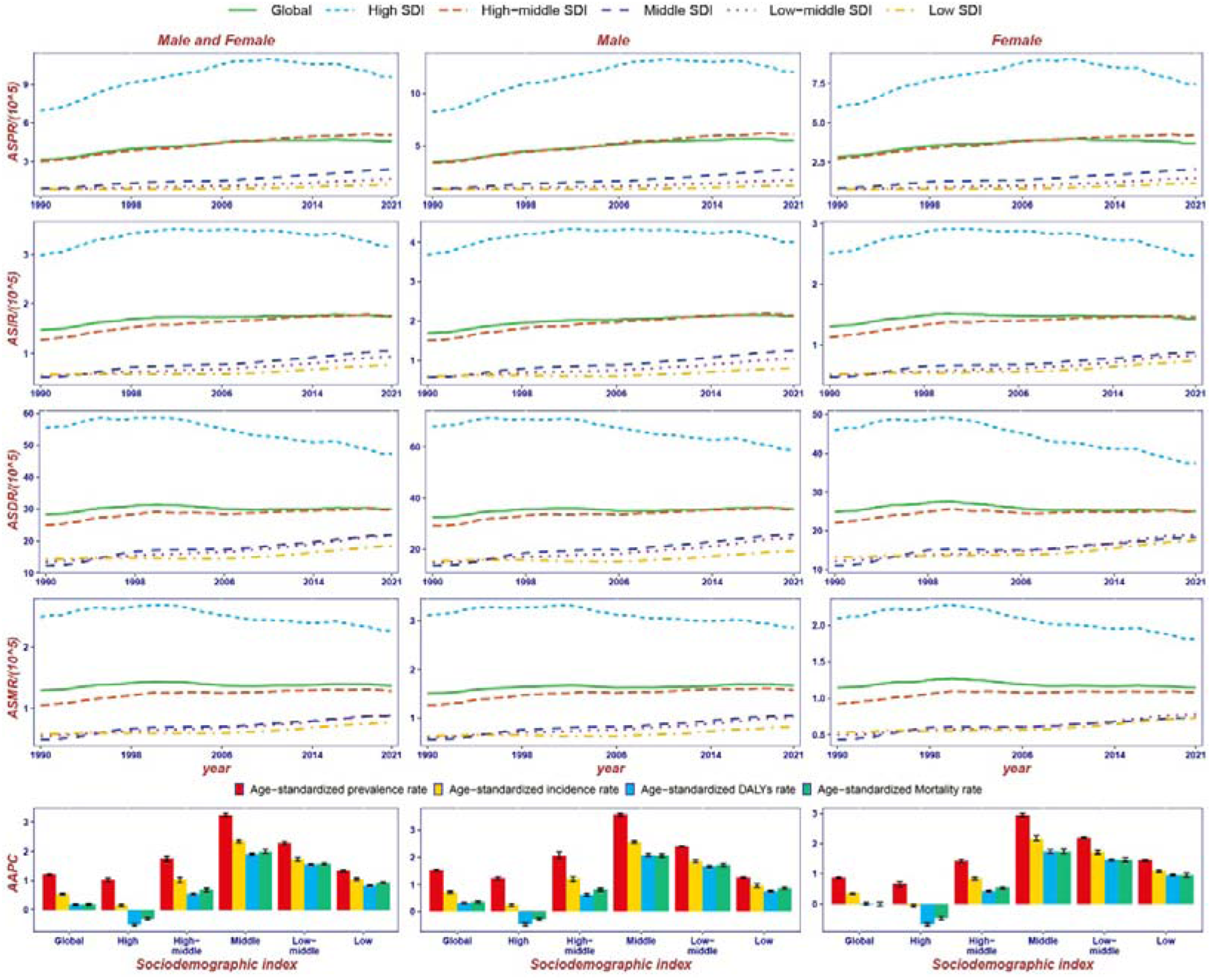
ASPR, ASIR, ASDR, and ASMR along with AAPC of MM from 1990 to 2021 in five SDI regions, categorized by sex.

### Risk factors associated with MM

In the GBD 2021, High BMI was reported as the only risk factor for MM, accounting for 7.96% of global MM ASDR and 7.84% of global MM ASMR (Figure S8A, B). Among the five SDI regions, the proportions of MM ASDR and ASMR due to High BMI both increased with SDI, reaching largest of 9.49% and 9.08% in high SDI regions, respectively. At the regional level, the proportions of ASDR and ASMR were the largest in High-income North America (11.52% and 11.05%) but the smallest in South Asia (3.61% and 3.35%). Both the proportions of MM ASDR and ASMR attributable to high BMI had increased from 1900 to 2021 in all regions, with the largest increase observed in East Asia. In addition, the proportion of MM ASMR and ASDR due to high BMI were both higher in females than in males. To further analyze the causal relationship between BMI and MM, we employed two-sample MR, which also confirmed that BMI was a risk factor for MM from a genetic perspective (Figure S9A). Moreover, other risk factors related to BMI, such as overweight, obesity class 1, obesity class 2, hip circumference, and waist circumference were all identified as significant casual risk factors for MM in the inverse variance weighted method with *P* < 0.05 (Figure S9B-F).

### Incidence rate predicted by BAPC model

According to the BAPC analysis, the incidence rate for males was predicted to decrease slightly from 2.08 in 2021 to 1.99 per 100,000 population in 2050, and for females, it was predicted to decrease from 1.69 to 1.34 per 100,000 population (Figure 4A, B). Nevertheless, the incidence rate will increase among individuals aged 20 to 69 whereas decrease among individuals aged 70 and older for both sexes, suggesting a potential trend of increasing MM burden in younger age groups (Figure 4C, D). Additionally, in the next 29 years, males will still have a higher incidence rate than females in the age range of over 20.

**Figure 4.**
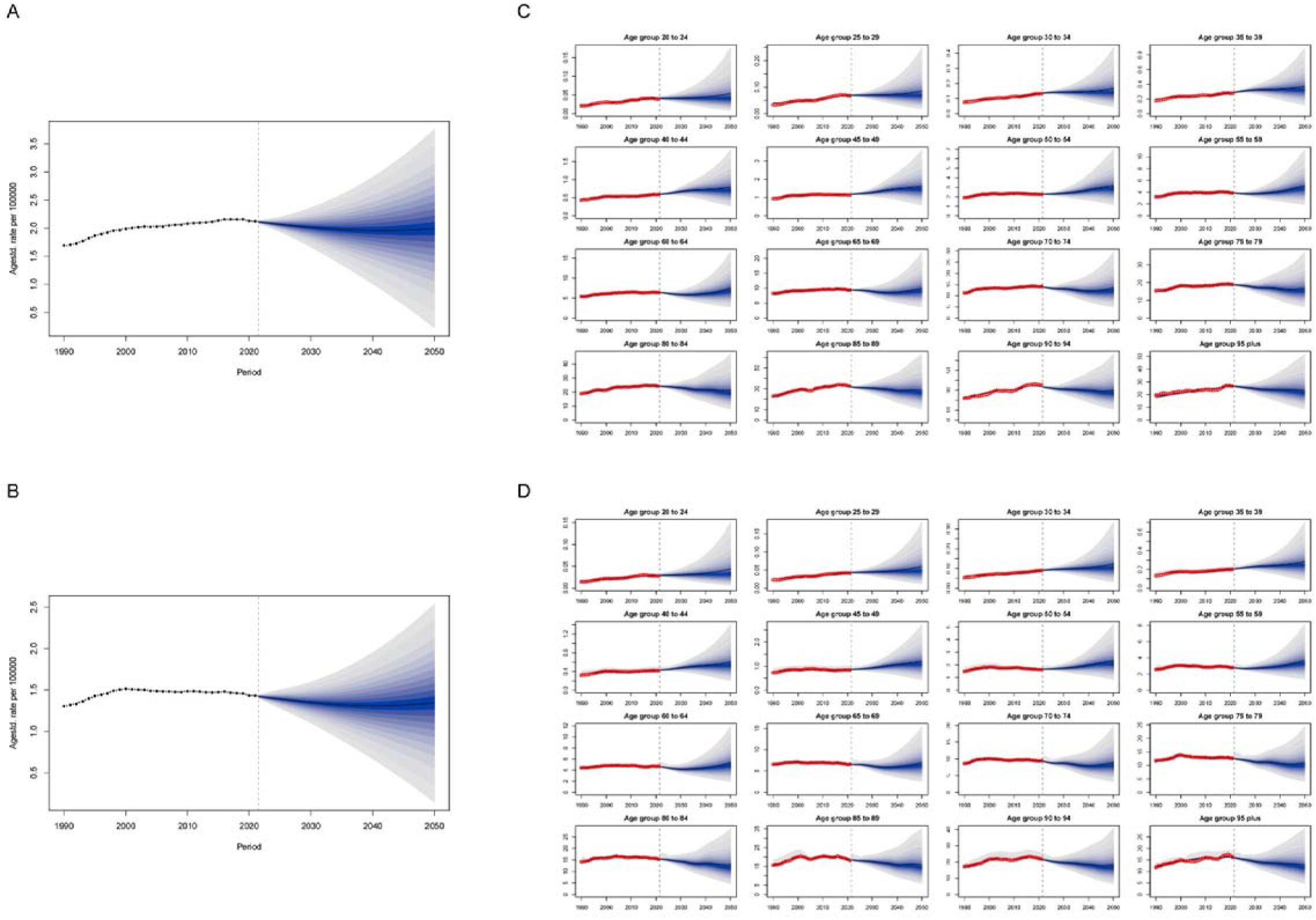
Trends of global MM incidence rate from 1990 to 2050 by sex and age groups. (A)The incidence rate of males across all ages; (B) The incidence rate of females across all ages; (C)The incidence rate of males in different age groups; (D) The incidence rate of females in different age groups.

## Discussion

Using the latest data from GBD 2021, this study is the most comprehensive evaluation on the growing global burden of MM. Our study revealed that over the past 31 years, the global burden of MM had consistently risen, posing significant challenges to global health systems. Meanwhile, the burden of MM varied across countries, sexes, and age cohorts, particularly heavy in males, people aged 50 years and older, and those from high SDI countries. Furthermore, we conducted a detailed analysis of BMI-related risk factors for MM. Finally, we predicted that the incidence rate of MM would decrease slightly by 2050.

All the ASRs were relatively high in Australasia and Western Europe. Countries with high SDI, such as New Zealand and Monaco had higher ASPR and ASIR while Mali with low SDI had the lowest ASPR and ASIR. These results were consistent with previous studies, indicating a higher hematological cancer burden in high-income countries.^6^ The significant disparities in the burden of MM may be attributed to differences in access to healthcare services and economic levels across countries. It had been reported that in high SDI countries, the rising incidence of disease was likely associated with better access to healthcare systems, which allowed for earlier detection and diagnosis.^19,20^ Conversely, in Mali and other low SDI African countries, the late diagnosis of MM had been attributed to inequalities in access to healthcare and inadequate health services^21,22^. In addition, it is noteworthy that among the five SDI regions, only the high SDI region experienced a decline in both ASDR and ASMR from 2004 to 2021. This may be attributed to the development of autologous hematopoietic stem cell transplantation and new drugs, such as bortezomib, which had extended the survival of patients.^23^ In recent years, the evolution of chimeric antigen receptor T cell technology further reduced the mortality rate of MM.^24^

Our study also found that the burden of MM was diverse in different age groups and sexes. In the elderly population, all ASRs are higher than in other age groups. This finding is consistent with the SEER database, which had reported that the incidence of MM increases with age.^25^ Existing studies have indicated that aging leads to a decline in stem cell regeneration, resulting in impaired physiological and immune functions, which makes the elderly more susceptible to cancer.^26,27^ Furthermore, once elderly individuals are diagnosed with MM, their poor tolerance to drug side effects impacts treatment efficacy, leading to higher mortality and DALY rates.^28^ In addition, males demonstrated higher ASRs compared to females, which may be partly due to the difference in lifestyles, such as smoking, drinking, and occupational exposure, including farming, nuke industry, chemical industry and medical radiation.^29^

The result of the two-sample MR analysis indicated that BMI and other obesity-related metrics, such as waist circumference, obesity class, and hip circumference, were significant risk factors with causal relationship to MM. In overweight populations, adipocytes can produce interleukin-6, which had been reported as an effective growth factor for MM.^30^ In addition, high insulin-like growth factor-1 in obese people had also been demonstrated to prevent apoptosis of MM cells and induce their proliferation.^31^ Overweight was a risk factor for various types of cancer which had gradually become a serious global threat.^32^ Public health strategies aimed at reducing obesity rates and promoting healthy lifestyles should play a crucial role in mitigating the burden of MM, particularly in regions where obesity is prevalent.

According to the BAPC model, the MM incidence rate is expected to decline among the elderly while rise in younger age groups. Compared to the elderly, middle-aged and younger individuals are more likely to experience obesity and be exposed to other risk factors, such as the chemical industry and medical radiation. This potential shift in the age distribution of MM highlighted the need for early detection and intervention. As stated in the WHO Rehabilitation 2030 initiative, ensuring equitable health services for MM patients globally and providing adequate resources for programs targeting the prevention and modification of risk factors are critically important on a global scale.^33,34^

This study has some limitations in assessing the burden of MM. First, due to underdeveloped cancer reporting mechanisms and infrastructure, the availability of data from certain countries and regions is relatively low. Additionally, since the GBD 2021 did not provide a classification of MM subtypes, more detailed studies were needed to evaluate the disease burden of different subtypes of MM.

## Conclusion

In conclusion, MM remained a significant and growing public health problem globally, particularly in regions with higher SDI levels. The burden of MM varied across different age groups and genders, being more severe among the elderly population and males. In addition, MR analysis revealed a causal relationship between MM and risk factors related to BMI, such as overweight, obesity class 1, obesity class 2, hip circumference, and waist circumference. By 2050, the incidence rate of MM was predicted to increase in the age group of 20-69 and remain higher in males than in females. More effective early detection strategies and strengthened healthcare infrastructure are keys to reducing the burden of MM.

## Data sharing

Original data generated and analyzed during this study are included in this published article or in the data repositories listed in References

## Acknowledgements

Yongshuai Jiang, Mingming Zhang, and Wenhua Lv designed this study. Chang Wang, Yan Guo, Ning Wang, and Qinduo Ren drafted the initial manuscript and finalized the manuscript with comments from all other authors. Yuan Xu, Songlin Lu, and Yuping Zou collected and analyzed the data. Ruilin Li, Jiacheng Wang, Linna Yuan, and Wei She validated the data and provide supervision. Hongsheng Tian, Shuo Bi, and Xuying Guo provided technical assistance. Ye Ma, Hongmei Sun, and Chen Zhang participated in the interpretation of the data. Yu Dong, Zhenwei Shang, and Hongchao Lv provided important comments on the manuscript. All authors reviewed the drafted manuscript for critical content and approved the final version of the manuscript. The corresponding author (Yongshuai Jiang) attests that all listed authors meet authorship criteria and that no others meeting the criteria have been omitted.

