## Supplementary appendix for "The burden trend and risk factors of multiple myeloma from 1990 to 2021: a systematic analysis for the Global Burden of Disease Study 2021"

### **Methods**

#### **Data**

The Global Burden of Disease, Injuries, and Risk Factors Study (GBD) 2021 was the most comprehensive global observational epidemiologic survey, which covered 288 causes of death, 371 diseases and injuries, and 88 risk factors in 204 countries and territories. GBD 2021 utilized data from vital registration, verbal autopsy, surveys, censuses, surveillance systems, and cancer registries, among others to estimate prevalence, incidence, mortality, years lived with disability (YLDs), years of life lost (YLLs), and disability-adjusted life-years (DALYs) across different age groups, sexes, geographical locations, time periods, and cause levels.^1^ All data of multiple myeloma (MM) used in this study were obtained from the Global Health Data Exchange (<https://vizhub.healthdata.org/gbd-results/>), including (1)global age-standardized and age-specific prevalence, incidence, mortality, DALY rates and cases by gender from 1990 to 2021; (2) sex-specific and age-standardized prevalence, incidence, mortality, DALY rates and cases across 21 regions and 204 countries and territories from 1990 to 2021; (3) regional sex-specific and age-standardized prevalence, incidence, mortality, DALY rates and cases by socio-demographic index (SDI); (4) the age-standardized mortality and DALY rate of MM attributable to high body mass index (BMI).

#### **Age-standardized rate**

Age-standardization refers to a statistical method of processing demographic data which could eliminate the impact of different age structures across populations and ensure the comparability of statistical indicators.^2^.The age-standardized rate (ASR) can be calculated using the following formula:^3^

$$\boldsymbol{ASR=}\frac{\sum_{\boldsymbol{i=1}}^{\boldsymbol{A}} \boldsymbol{a}_{\boldsymbol{i}}\boldsymbol{w}^{\boldsymbol{i}}}{\sum_{\boldsymbol{i=1}}^{\boldsymbol{A}} \boldsymbol{w}_{\boldsymbol{i}}}$$

where $\boldsymbol{A}$ is the upper limit of age, $\boldsymbol{i}$ represents the $\boldsymbol{i}^{\boldsymbol{th}}$ age group, $\boldsymbol{a}_{\boldsymbol{i}}$ is the age specific rate, $\boldsymbol{w}_{\boldsymbol{i}}$ denotes the weight of the $\boldsymbol{i}^{\boldsymbol{th}}$ age group in the standard population from GBD 2021. The application of ASRs allows for more accurate assessment of the disease burden of MM among countries or regions in different periods with different population age structures.

#### **Two-sample Mendelian randomization**

Mendelian randomization (MR) is a statistical method that uses genetic variation an instrumental variable (IV) to address causal questions regarding whether modifiable exposures influence health, developmental, or social outcomes.^4^ MR is based on the three key assumptions: (1) a stable and strong association between the IV and the exposure; (2) independence between the IV and confounding factors that affect both the exposure and the outcome; (3) on causal pathways between the IV and the outcome, other than through the exposure.^5^ Using the above methods, MR avoids bias caused by unobserved confounding between exposure and outcome and provided a rapid and reliable approach for assessing causal relationships. Considering that the GBD 2021 report identifies high BMI as a risk factor for MM, we employed two-sample MR to further investigate the causal relationships between weight-related factors, including BMI, overweight, obesity class 1, obesity class 2, hip circumference, and waist circumference. In MR analysis, the Inverse Variance Weighting (IVW) method was used as the primary analytical approach. The genetic variation data for the exposures and MM were sourced from the genome-wide association study (GWAS) summary data of the OpenGWAS project (https://gwas.mrcieu.ac.uk).

### **Supplementary Figures**

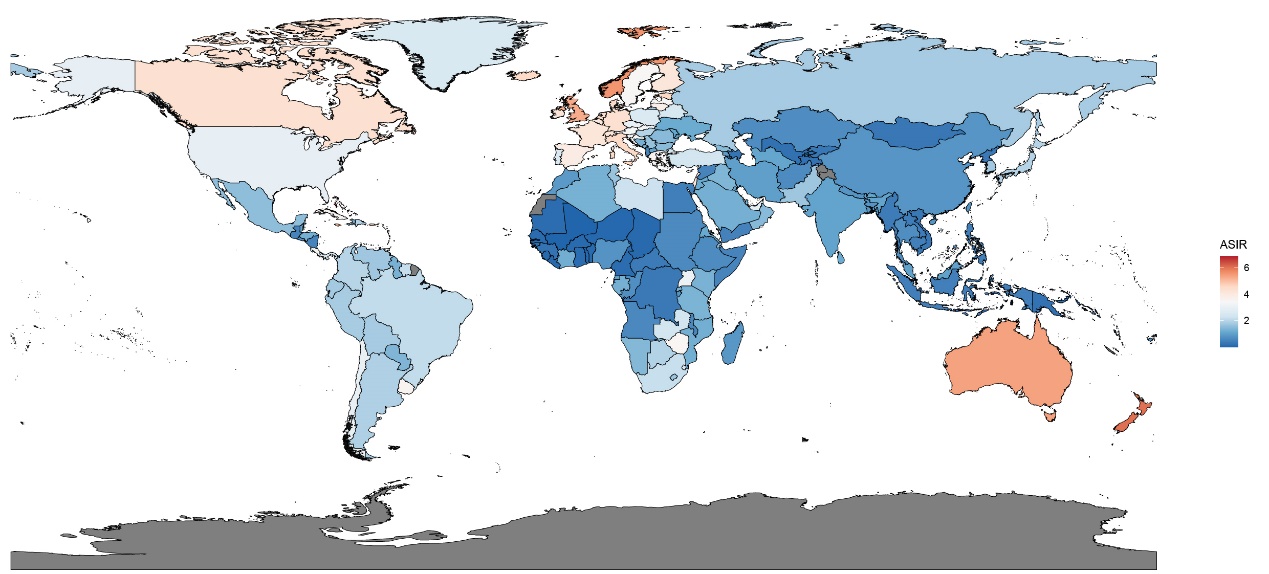

Figure S1. ASIR of MM in 204 countries and territories in 2021.

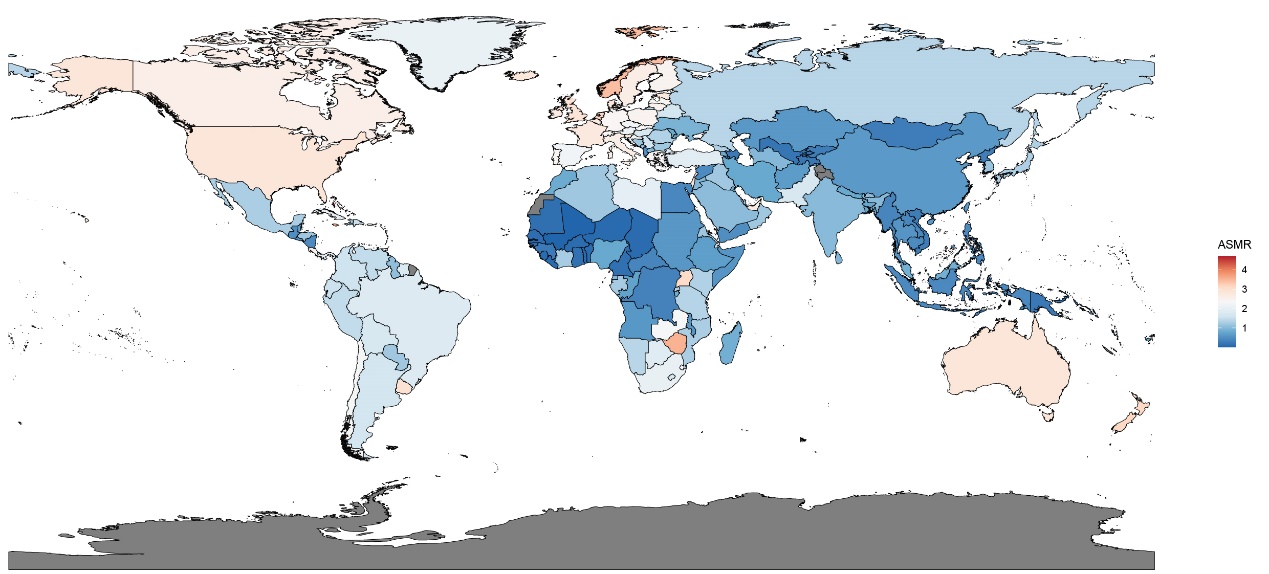

Figure S2. ASMR of MM in 204 countries and territories in 2021.

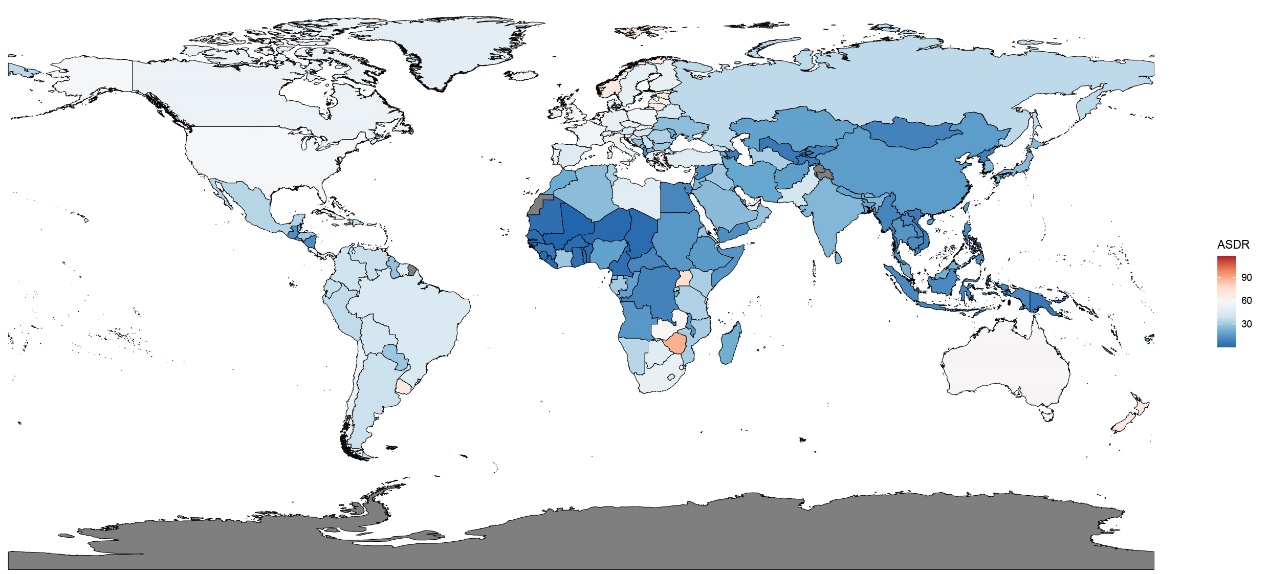

Figure S3. ASDR of MM in 204 countries and territories in 2021.

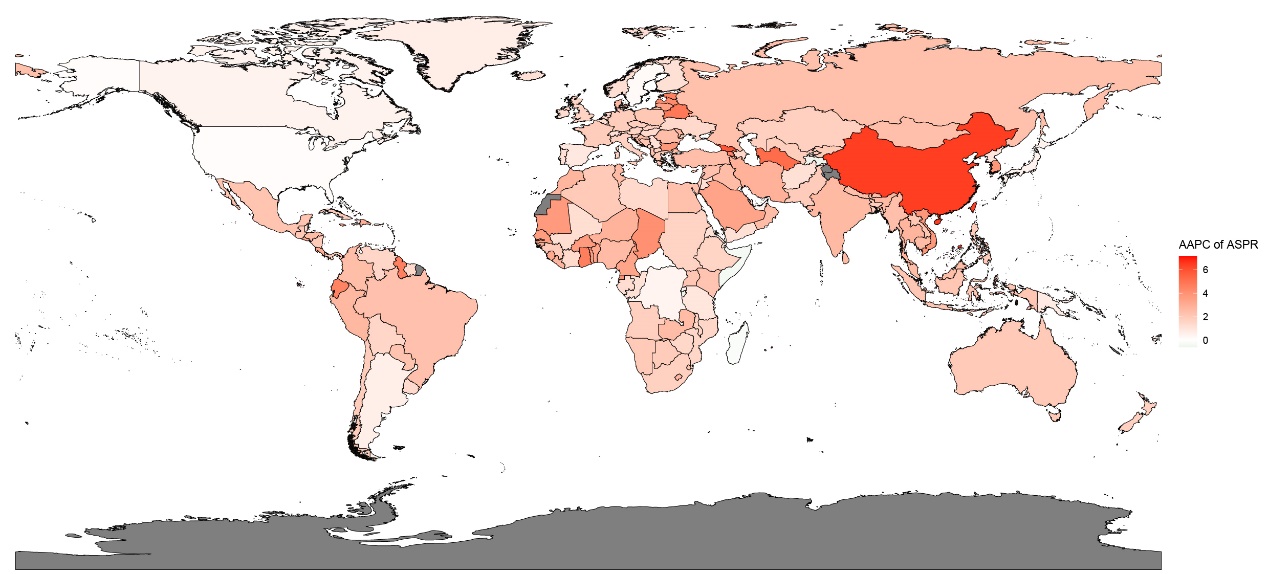

Figure S4. AAPC of ASPR for MM in 204 countries and territories in 2021.

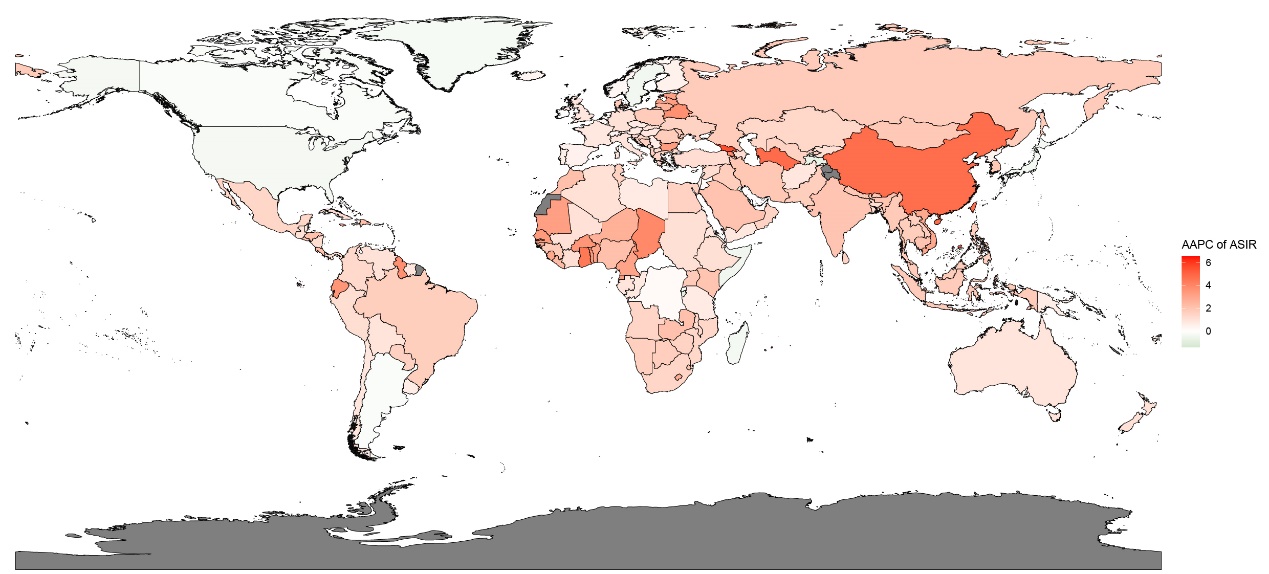

Figure S5. AAPC of ASIR for MM in 204 countries and territories in 2021.

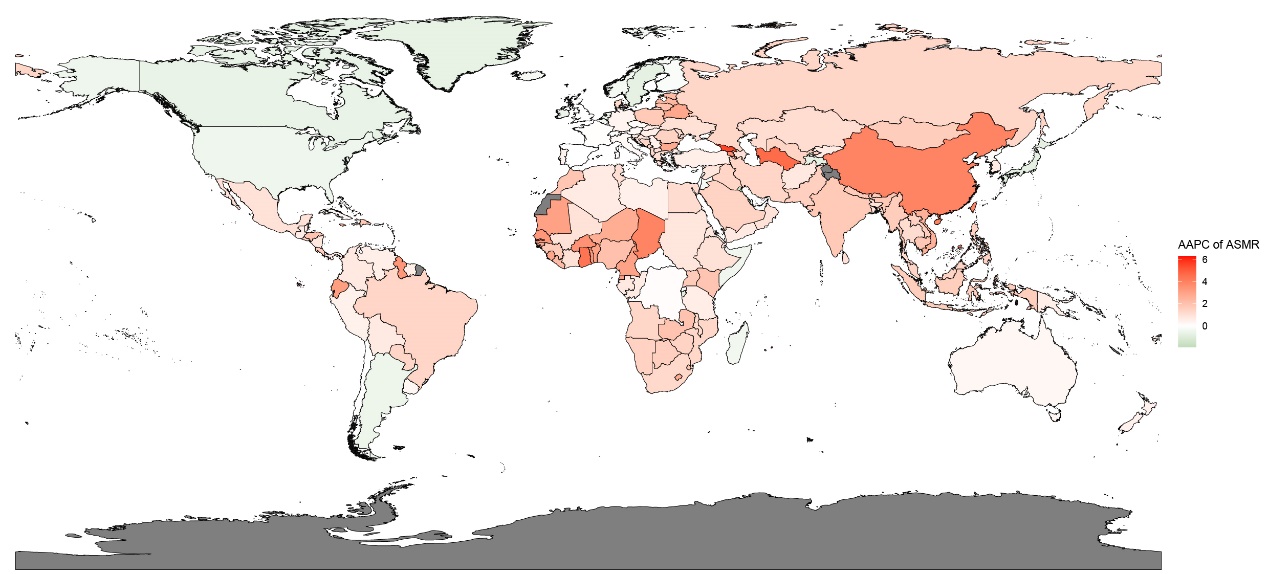

Figure S6. AAPC of ASMR for MM in 204 countries and territories in 2021.

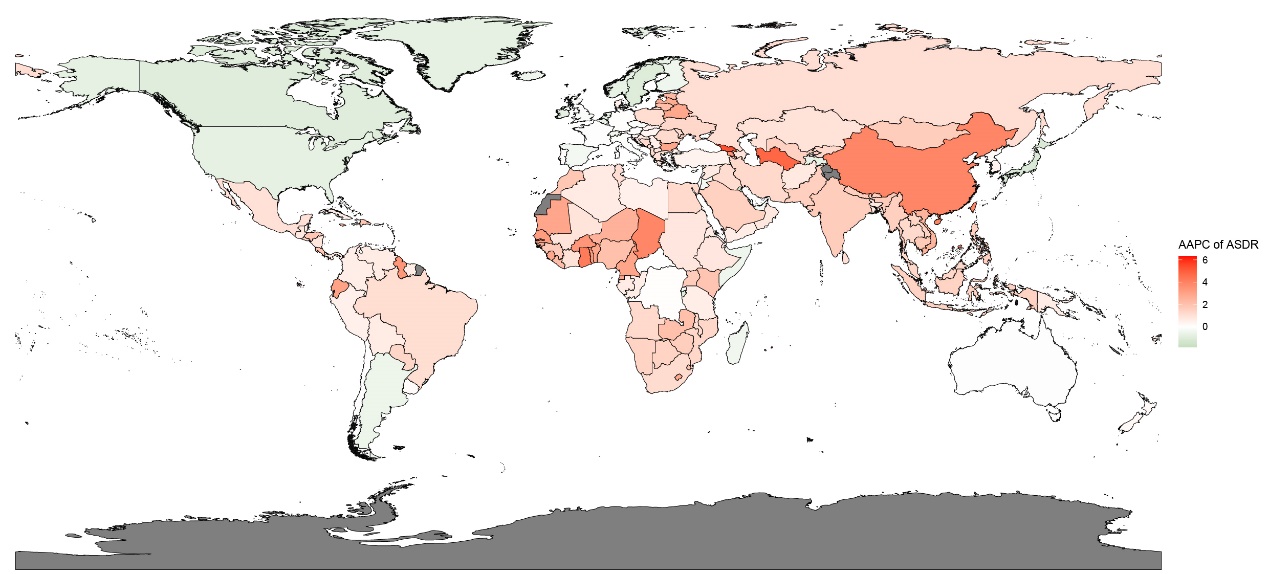

Figure S7. AAPC of ASDR for MM in 204 countries and territories in 2021.

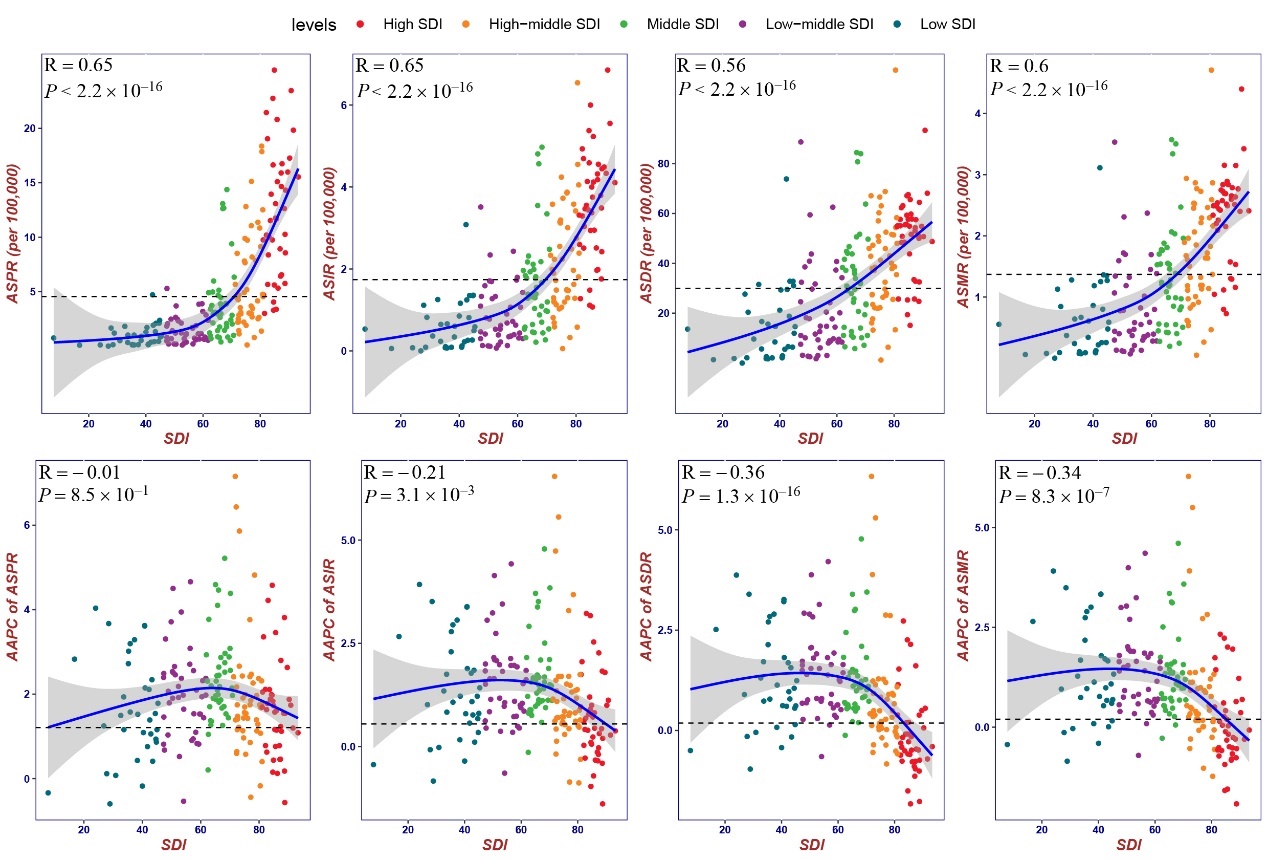

**Figure 4. The relationship between SDI and ASRs of MM in 2021, as well as corresponding AAPC from 1990 to 2021. The dashed line represented the global ASR level.**

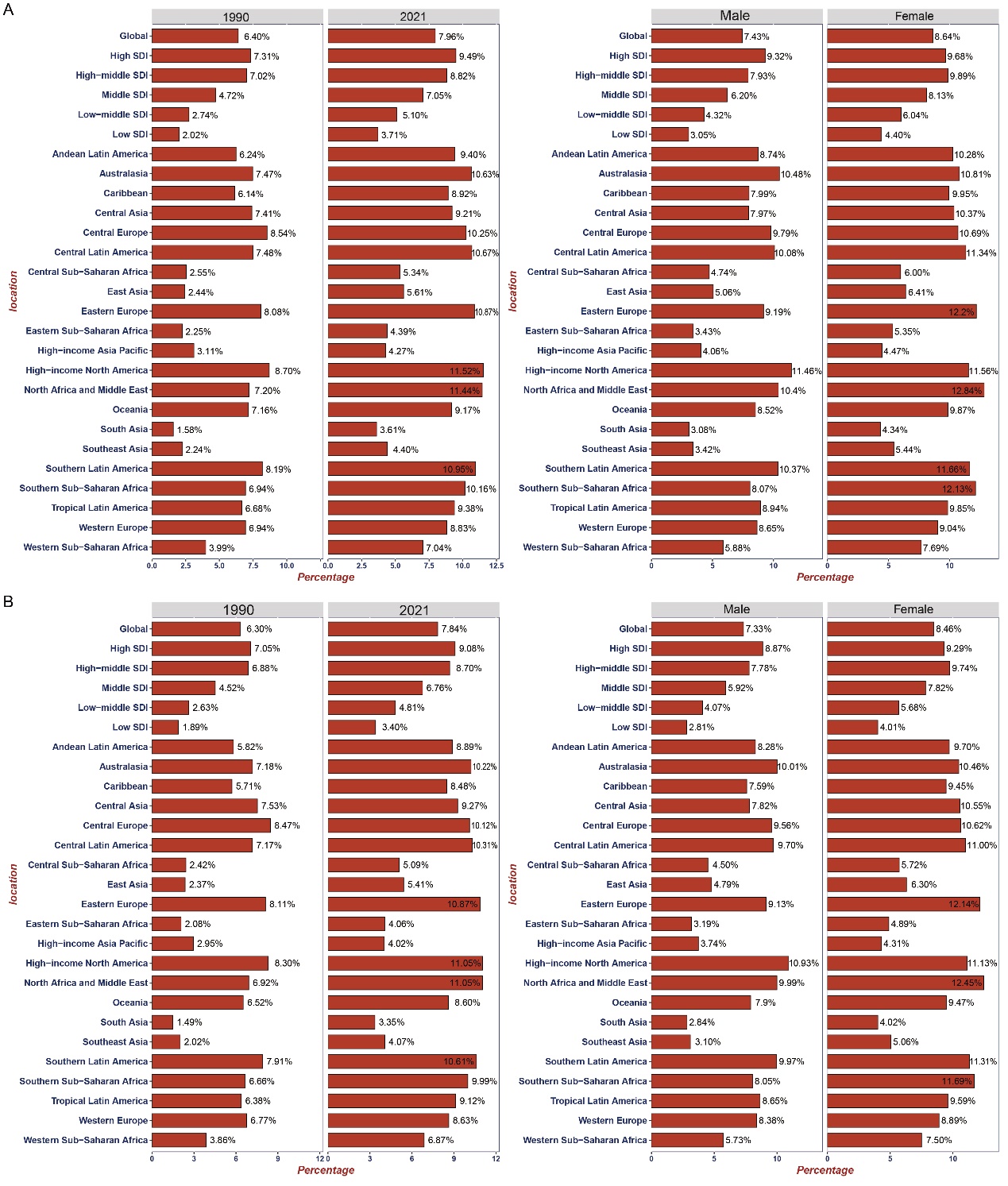

**Figure 5. Proportion of MM ASDR and ASMR attributable to high BMI, classified by year and sex. (A)ASDR; (B) ASMR.**

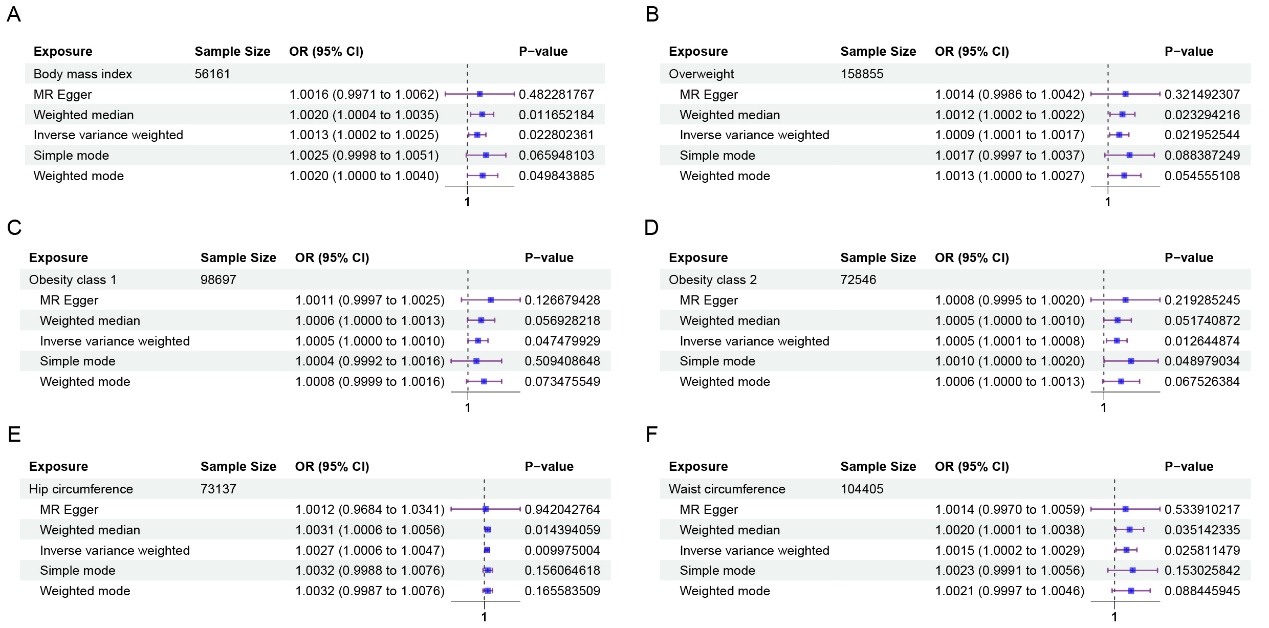

**Figure 6. Forest plot of BMI-related risk factors with causal relationships to MM. (A) Body mass index; (B) Overweight; (C) Obesity class 1; (D) Obesity class 2; (E) Hip circumference; (F) Waist circumference.**

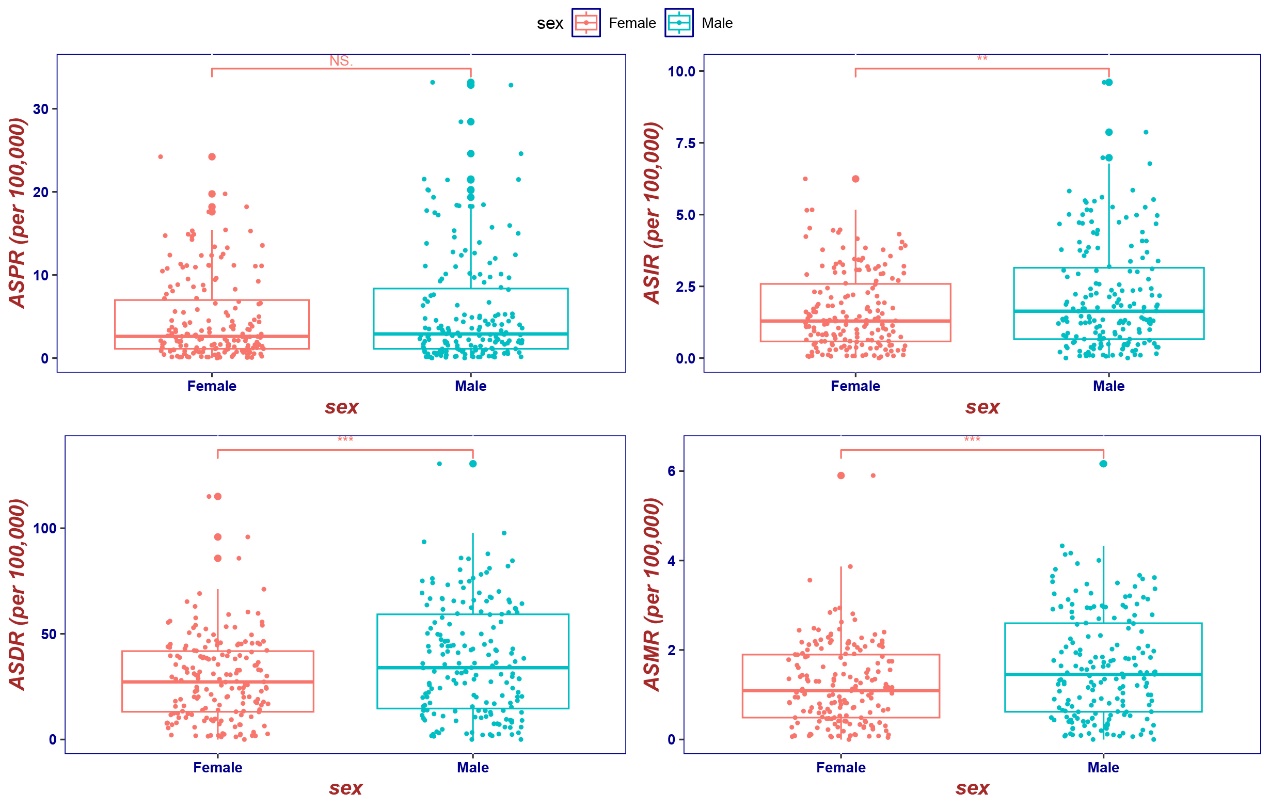

Figure S8. Sex Differences in ASRs of MM in 2021 by 204 countries and territories.
